# Ocular toxicity and Hydroxychloroquine: A Rapid Meta-Analysis

**DOI:** 10.1101/2020.04.28.20083378

**Authors:** Matthew Michelson, Tiffany Chow, Neil Martin, Mike Ross, Amelia Tee, Steven Minton

## Abstract

Rapid access to evidence is crucial in times of evolving clinical crisis. To that end, we propose a novel mechanism to answer clinical queries: Rapid Meta-Analysis (RMA). Unlike traditional meta-analysis, RMA balances quick time-to-production with reasonable data quality assurances, leveraging Artificial Intelligence to strike this balance. This article presents an example RMA to a currently relevant clinical question: *Is ocular toxicity and vision compromise a side effect with hydroxychloroquine therapy?*

As of this writing, hydroxychloroquine is a leading candidate in the treatment of COVID-19. By combining AI with human analysis, our RMA identified 11 studies looking at ocular toxicity as a side effect and estimated the incidence to be 3.4% (95% CI: 1.11-9.96%). The heterogeneity across the individual study findings was high, and interpretation of the result should take this into account. Importantly, this RMA, from search to screen to analysis, took less than 30 minutes to produce.

## Introduction

The COVID-19 pandemic caused by the Severe Acute Respiratory Syndrome Coronavirus 2 (SARS-Cov-2) presents clinicians and regulators with multiple challenges, including access to timely and crucial information. To mitigate this issue, many segments of the healthcare industry have looked to Artificial Intelligence (AI) for support.

The capacity of AI to aggregate and process massive volumes of information is emerging as particularly crucial in the current moment, especially as so much data is available it can be overwhelming for humans to evaluate.^1^ AI technology, such as that from Evid Science^a^ can unburden some of this overload by automatically processing the written text of medical papers, turning the text into a more consumable, structured set of data that can be easily searched and analyzed. Essentially, the AI turns all of the written articles into spreadsheets of results.

Meta-analysis and Systematic Literature Review are the gold standards for evidence,^3^ but they take significant time and effort to produce (often as long as a year^4^) and are therefore rarely updated.^5,6^

To produce results in a more timely manner, in this paper we propose the Rapid Meta-Analysis (RMA). An RMA follows the same general framework methodology of a Meta-Analysis, but leverages technology at every step, yielding a much faster time-to-production. Some data quality may be compromised due to the emphasis on fast time-to-production, but the ability to generate answers so quickly may warrant this trade off.

This article introduces RMA by way of practical example. Hydroxychloroquine (HCQ) has been available since the 1950s^2^ and has been used to treat malaria, lupus erythematosus, and rheumatoid arthritis. Most recently, has been highlighted as a potential intervention to support patients with Covid-19. While the efficacy outcomes of hydroxychloroquine are different in each clinical condition for which it is used, adverse events tend to be consistent. In this study, we use RMA to answer a specific clinical question regarding HCQ and the degree to which ocular toxicity is a side effect. This is an important clinical question, yet the authors could not find a suitable aggregation of results.

Therefore, we leveraged the Evid Science clinical-outcomes database to produce an RMA in order to find relevant studies, screen to those focused on HCQ and vision issues, and then perform the meta-analysis computation. The data for the RMA is provided in Table 1. Based on results from 11 studies (N = 3,585), we expect to see major eye issues 3.4% of the time (95% CI: 1.11-9.96%). We note high heterogeneity across the studies (I^2^ is 97%) so we caution when interpreting these results. Importantly, the entire process from search to analysis took less than 30 minutes.

**Table 1:**
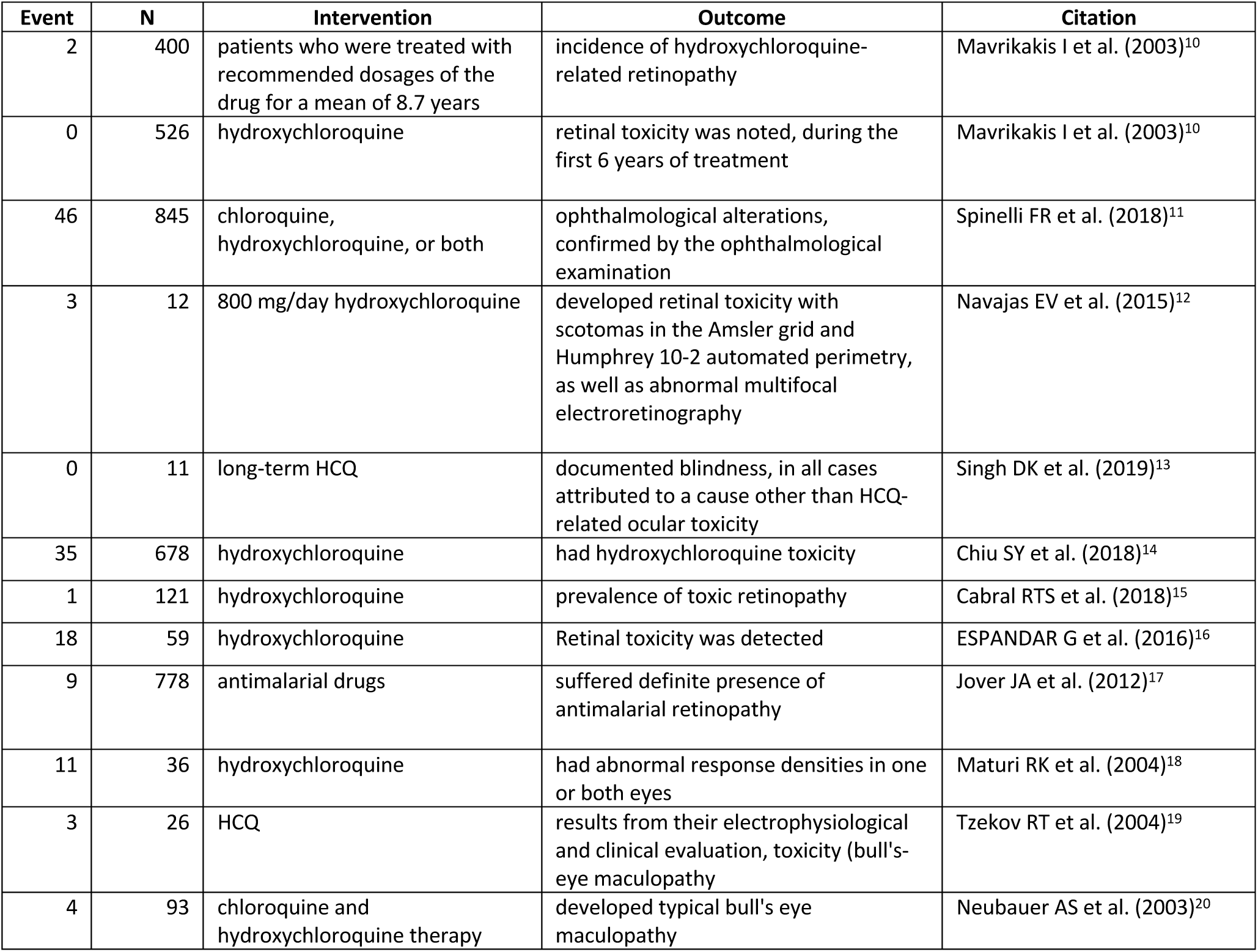
Results from Included papers.

To be clear, a RMA, such as this, is meant to raise awareness, and should not be treated as a full Systematic Literature Review and Meta-Analysis that should strictly guide treatment. The data and results are current as of 4-11-2020.

## Background

The core innovation of an RMA is replacing as many of the steps as possible with machine intelligence, as has been proposed previously.^7,8^ Machines are not yet at the point where they can simply provide an answer to a posed question, so RMA instead replaces as many manual steps as possible with machine-assisted (or entirely AI) steps. The goal is that each step could eventually be replaced with AI.

To make this idea concrete, consider Figure 1 below. On the left of Figure 1, we see the standard steps (at a high level) for meta-analysis. On the right is there equivalent technology replacement.

**Figure 1:**
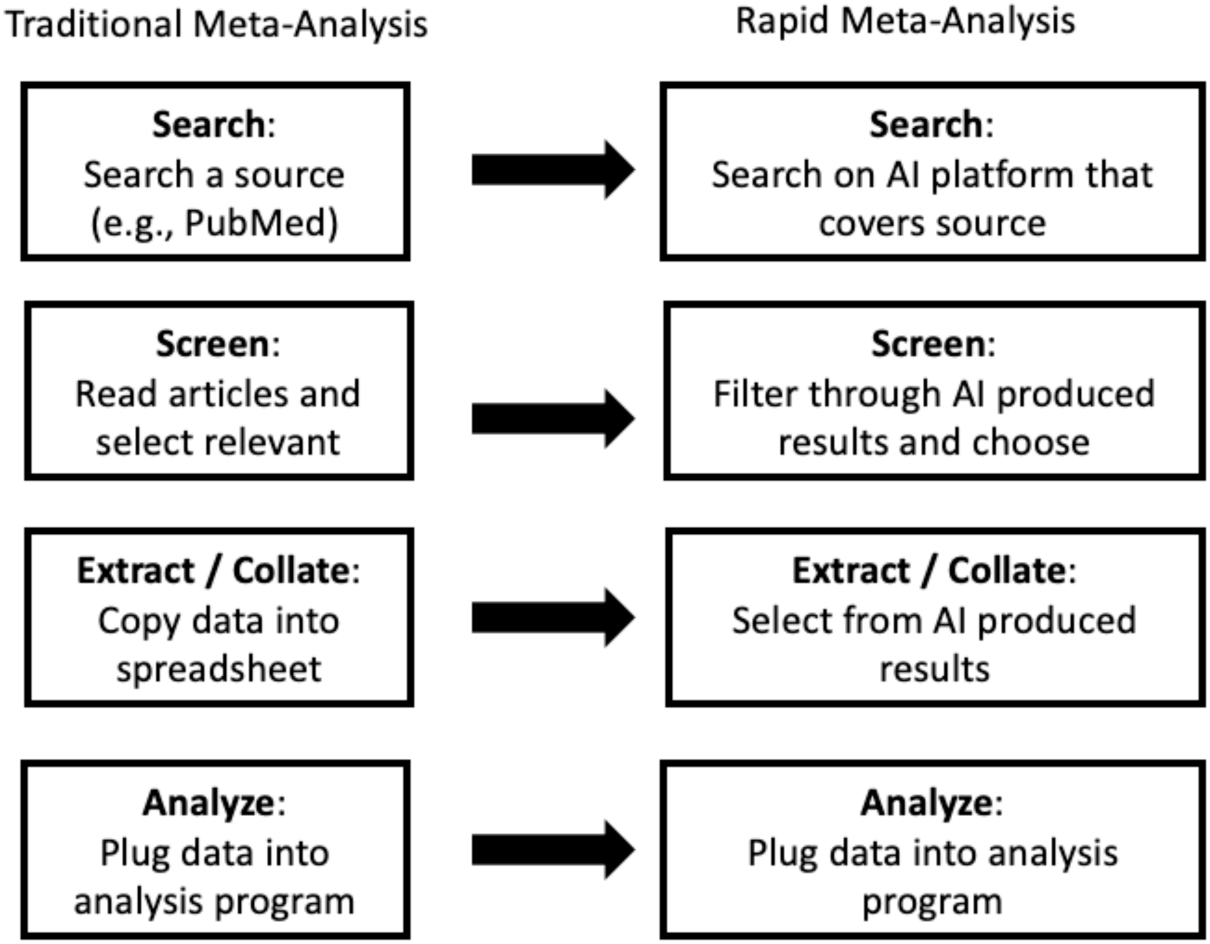
Traditional meta-analysis (simplified, left) vs. RMA using AI (right)

For this RMA, we leverage the Evid Science clinical-outcomes database for searching and screening (though any suitable AI system could provide similar benefit). This database was built using the Evid Science AI, which is capable of turning written text about results into a “structured” representation (e.g., a row in a database or spreadsheet).

Consider Figure 2 below. It shows a sentence about toxicity detected for a set of patients. The AI is able to break this sentence down into fields (such as result, intervention and outcome), automatically. It knows that 18 is a number of patients, and since that represents 30.5% of the patients, it must be 18 of 59. It also knows that the 18 was associated with “Retinal toxicity being detected”, in contrast to the 5, which is associated with “color vision impairments.”

**Figure 2:**
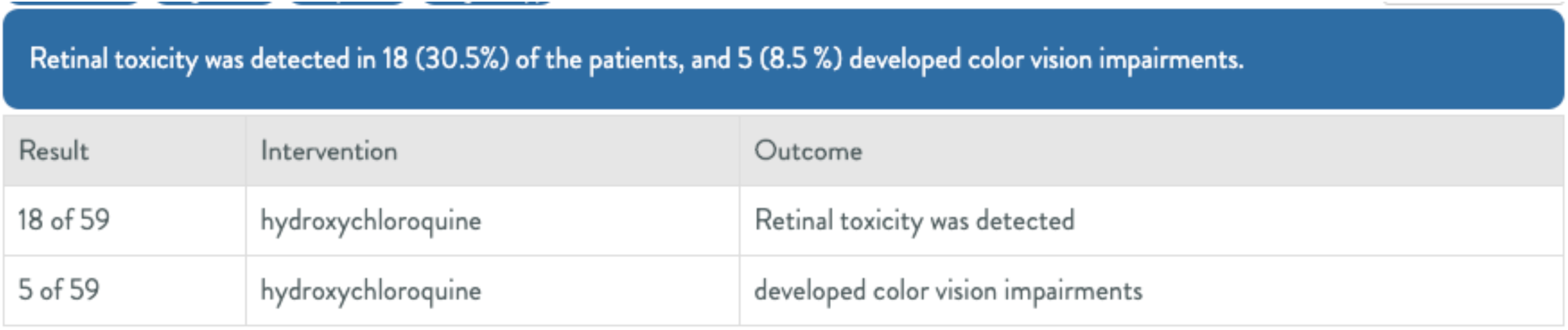
An example of AI generated results from text.

For the AI to learn to perform this task, researchers at Evid Science employed Supervised Machine Learning. In this methodology, the researchers initially gave the AI very explicit examples of the type of output they want (similar in format to Figure 2 - they are sets of sentences and the associated structured results).

After developing a data set of thousands of such examples, from across varied articles in the literature, the machine learns to produce these types of output for brand new sentences. To be clear, the articles chosen for training are selected from multiple disease topics and with various interventions – not just focused on HCQ. As the system improves, it can even be taught to correct mistakes, rather than having to start with fresh examples each time, thereby limiting the effort involved in refining its learning.

In previous work, we demonstrated a similar process to RMA, using the Evid Science AI to replicate the results from a Systematic Literature Review.^9^ Crucially, by leveraging AI we produced the results in six days, rather than the months it took to produce the original. Also, given the time between the original publication and AI-assisted version, 22 new relevant results had been published. We note, the current version of the AI used for this RMA is significantly more powerful than the version previously used for SLR replication.

The Evid Science clinical outcomes database used in this RMA is the result of running the AI over the entirety of the publicly (freely) available medical literature (PubMed). The current database has nearly 70,000,000 “facts” associated with results from articles which users can search and screen through.

Our RMA proceeds by searching and screening through this database, as we describe below. We note, the search itself (Step 1), leverages PubMed APIs, and therefore returns equivalent articles to PubMed.

Screening is then simplified, since the AI has processed the text into structured records that can be filtered and screened efficiently. For instance, we can simply filter to results associated with “toxicity” in the outcome (or other outcomes we care about). This is more efficient than manually reading each returned abstract, since one only screens articles in the filtered set.

After searching and screening, a user has the final data set for analysis. One can do many analyses directly within the Evid Science Web-based tool or export data to Excel and then analyze it with other programs (as we did).

## Methods

In this section we describe our RMA process, focusing on eye issues and HCQ.

To start, we did a search for hydroxychloroquine on the Evid Science platform^b^, and filtered down to results where the outcome discussed major vision impairments (e.g., “maculopathy,” “blind,” “toxicity” etc.). This yielded 22 candidate articles (from a possible set of 5,010 articles related to HCQ, of which 1,352 were identified as primary studies - e.g., trial or observational - by our AI and therefore included as possible articles to process results from).

After screening, we were left with 11 papers for our RMA. (The 11 excluded articles were published before 2000 or focused on diagnosis of ocular issues). The search took less than 1 minute and the screening took 22 minutes (most of the work was selecting the papers and lightly cleaning the results to make the table of results easier to read.) The results from search and screening are in Table 1 below. This is the input to our meta-analysis computation.

## Results

We then meta-analyzed the above results, using a generalized linear mixed model (in the R programming language), as these are binary occurrences of having the eye issue. We chose the Random Effects model results, and back traced the effect to compute the expected number of observations of the issue per 100 observations. We report this number and the confidence interval (3.4%; 95% CI: 1.11-9.96%). The code for this analysis was already written, so plugging in the data (Table 1) and running it took roughly 2 minutes (export data to excel, rename and select columns to conform to R-code input, run the code).

The forest plot of the meta-analysis is shown in Figure 3 below. Clearly there is heterogeneity (I^2^ is 97%), so these results warrant deeper inspection and cautious interpretation. The funnel plots of the results is shown in Figure 4.

**Figure 3:**
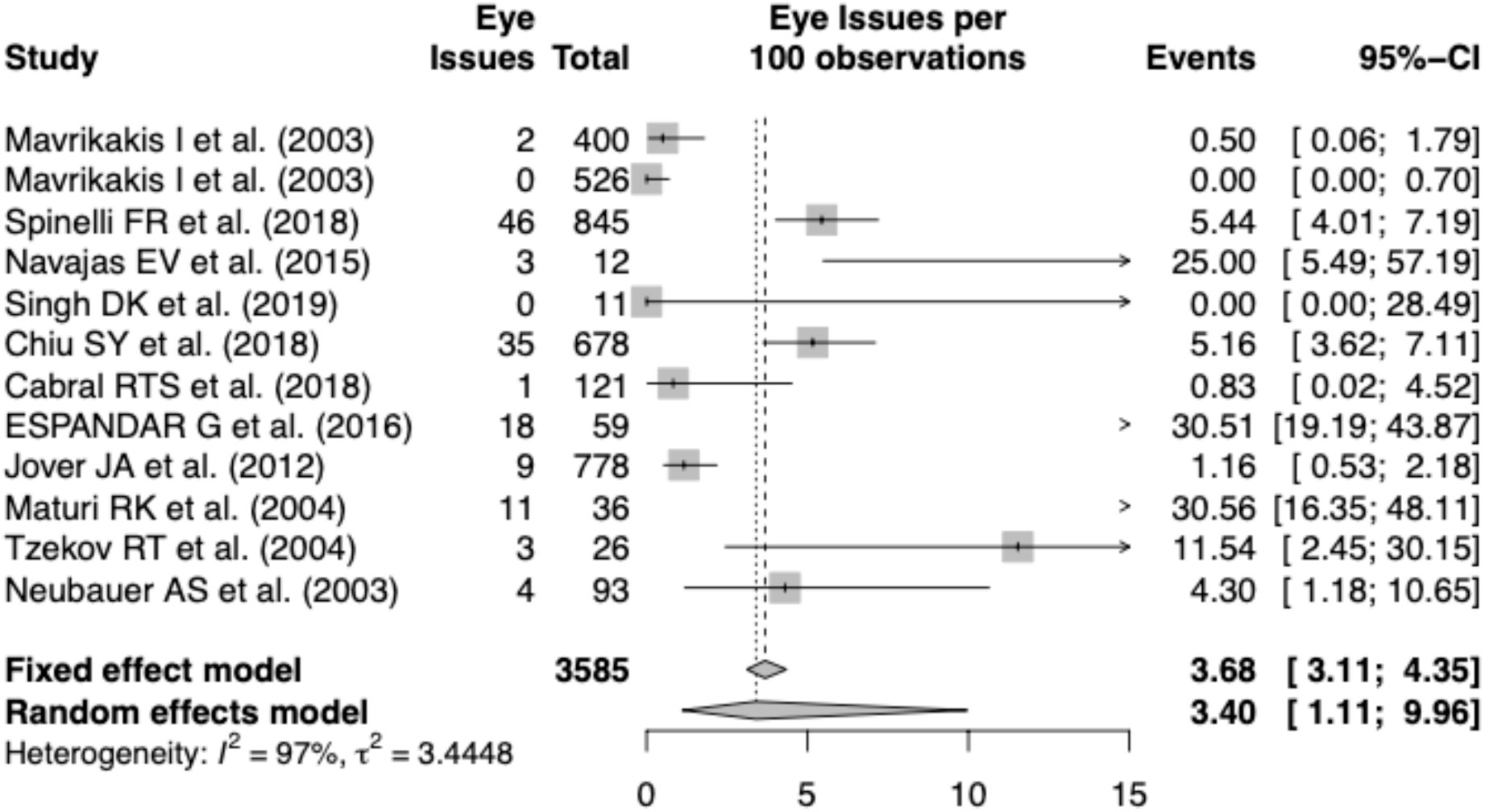
Forest plot for our RMA.

**Figure 4:**
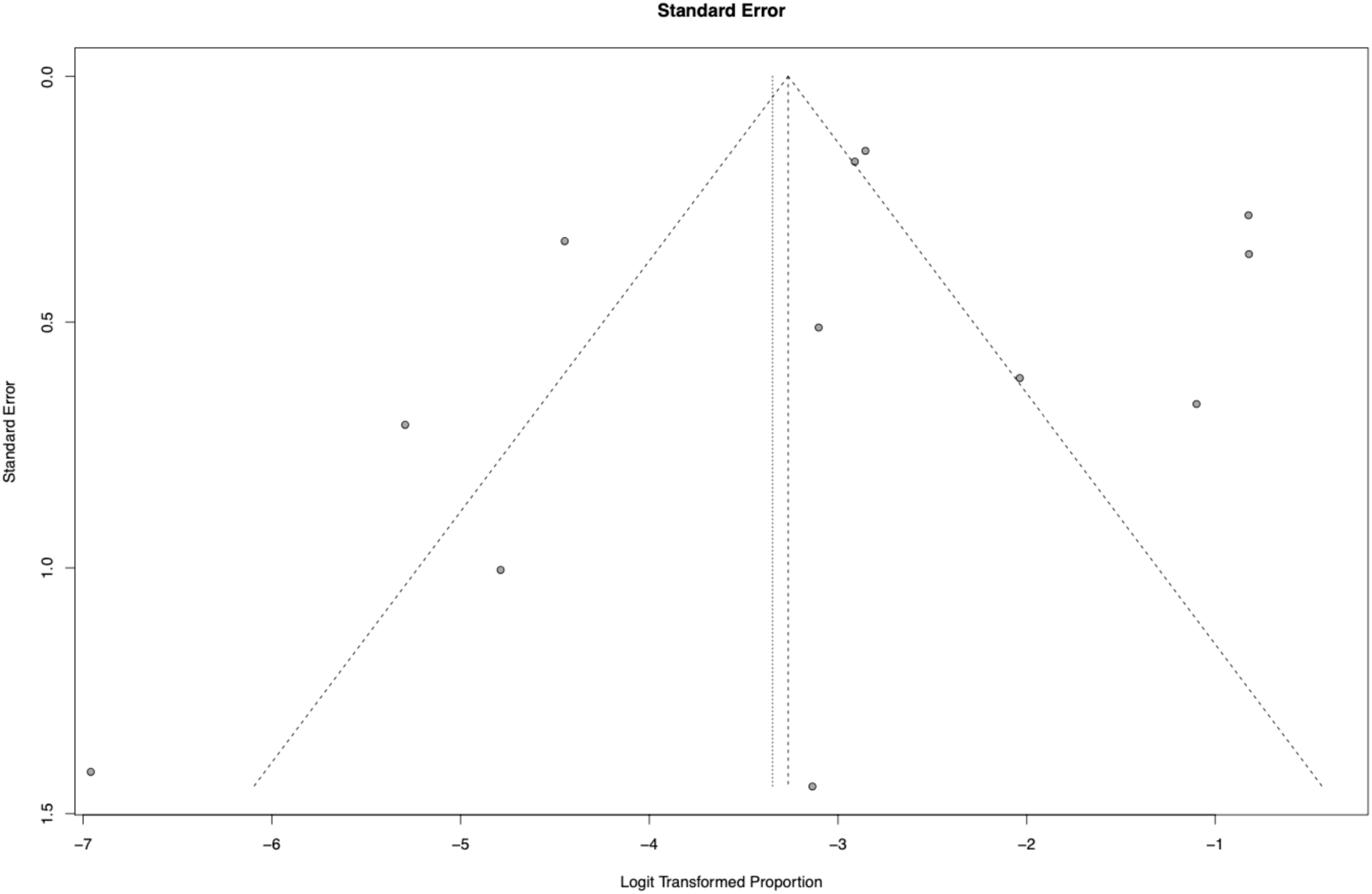
Funnel plot for our RMA.

Each step of our RMA, its output and its timing are shown below in Figure 5. This is RMA.

**Figure 5:**
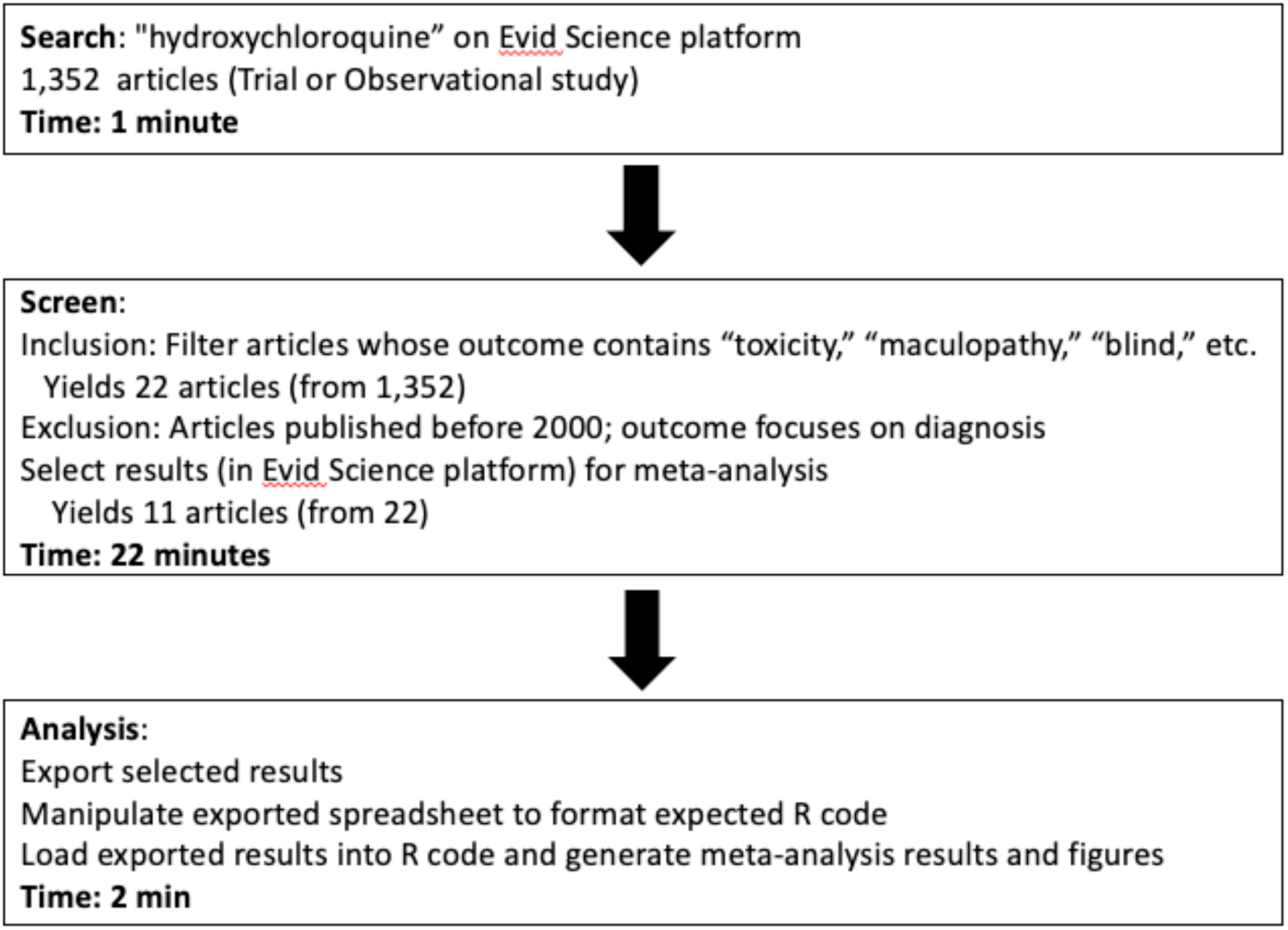
RMA for ocular toxicity associated with HCQ.

## Conclusion

In this article we present a new framework for answering clinical questions when time is at a premium and can be traded off for data quality and depth. We call this a Rapid Meta-Analysis and we demonstrated its utility in answering a clinical question about a drug being proposed to treat COVID-19. While the results raise further questions that need to be considered (e.g., regarding the high heterogeneity), they do bring attention to an important clinical issue with the drug hydroxychloroquine. And importantly, the whole assessment was done in less than 30 minutes.

## Data Availability

We include all of the data for the meta-analysis within a table in the paper for completeness and easy availability.

## Footnotes

a MM and TC are both employees of Evid Science

b In this study, we focus solely on PubMed abstracts, since those are freely available.

## Notes

### Competing Interest Statement

Matthew Michelson, Tiffany Chow, Mike Ross and Amelia Tee are employees of Evid Science, whose AI and data set were used in this publication.

### Funding Statement

No external funding

